# Does Practice Match Protocol? Outcomes of More-Acute Emergency Department Patients Seen After Less-Acute Patients Arriving Nearly-Simultaneously

**DOI:** 10.1101/2024.10.17.24315545

**Authors:** Temesgen Tsige, Rida Nasir, Daisy Puca, Kevin Charles, Sandhya LoGalbo, Lisa Iyeke, Lindsay Jordan, Melva Morales Sierra, David Silver, Mark Richman

**Author notes:** Author Contributions: Mark Richman, David Silver, Lisa Iyeke, Lindsay Jordan, Rida Nasir, Kevin Charles, Daisy Puca and Melva Morales Sierra: Conceived of the project and did the initial manuscript draft and literature search. Mark Richman, Temesgen Tsige and Sandhya LoGalbo: Created data collection tool, performed data collection and entry, integrated those findings and references into the manuscript, reviewed and edited final manuscript. Patient’s Consent: N/A. Trial/Systematic Review Registry: N/A.

## Abstract

**INTRODUCTION:** Emergency Departments (EDs) serve as the first line of healthcare, addressing a range of conditions from minor to life-threatening. With rising patient volumes, many EDs have adopted innovative models like the split-flow system to increase efficiency and manage overcrowding. The split-flow model, designed to improve efficiency, directs lower-acuity patients to a Fast Track area while higher-acuity cases remain in the main ED. However, this model can lead to lower-acuity patients being seen by a provider before more-acute patients, raising concerns about delayed care for more-acute patients. This study aims to investigate whether prioritizing less-acute patients impacts disposition outcomes for more-acute patients, focusing on admission rates and 30- and 90-day return visits. We hypothesized that more-acute patients seen after near-simultaneously arriving less-acute patients will experience higher rates of return visits and increased hospital readmissions compared to those seen prior to near-simultaneously arriving less-acute patients.

**MATERIALS AND METHODS:** This retrospective observational study assessed the impact of triage practices in the Emergency Department (ED) at Long Island Jewish Medical Center (LIJ), a 583-bed tertiary-care hospital. Adult patients (≥18 years) presenting to the ED between April 24 and December 13, 2023, were included, with a sample of 126 patient pairs triaged within 10 minutes of each other. Patients were categorized as high-acuity (ESI 1-2) or low-acuity (ESI 3-5). Data was drawn from electronic health records, including the Emergency Severity Index (ESI) level, the ED location to which the patient was triaged (either Fast Track vs. acute care area), return visits at 30 and 90 days, and patient disposition during the 30-day revisit.

**RESULTS:** The study comprised 126 patient pairs (252 patients in total). Overall, there were no statistically-significant differences in dispositional outcomes for more-acute patients based on the order in which they were seen. However, when more-acute patients were seen first by a physician, the results indicated that 10% returned to the ED within 30 days, with 63% requiring hospital admission. Additionally, 20% of these patients returned within 90 days. Conversely, when less-acute patients were prioritized, 22% of more-acute patients returned to the ED within 30 days, with only 45% requiring admission, and the 90-day return rate increased to 26%.

**CONCLUSION:** Research has demonstrated a connection between timely care and improved patient outcomes. Our previous study revealed that less-acute patients were seen prior to near-simultaneously-arriving more-acute patients approximately 40% of the time. This raises significant ethical concerns, as it contradicts the fundamental principle of emergency medicine, which emphasizes treating patients based on acuity. Although this current study found no significant differences in dispositional outcomes based on the order in which more-acute patients were seen, the trends suggest that seeing more-acute patients later might lead to worse outcomes. When more-acute patients were seen first, 10% returned to the ED within 30 days, with 63% requiring hospital admission, and 20% of indexed patients returned within 90 days. However, when less-acute patients were seen first, the 30-day return rate for more-acute patients increased to 22%, with 45% needing hospital admission, and the 90-day return rate rose to 26%. EDs must develop strategies to balance operational efficiency with the need to prioritize higher-acuity patients to ensure that operational practices do not compromise patient safety.

## INTRODUCTION

Emergency Departments (EDs) are the frontline of healthcare, handling everything from minor illnesses to life-threatening conditions. As patient volumes continue to rise and healthcare systems are pressed to do more with less, EDs have adopted innovative models like the split-flow system to increase efficiency and manage overcrowding^1-3^. The goal is to move patients through the system faster by directing those with less severe conditions (lower acuity) to a Fast Track area for faster care, while reserving the main ED for patients in critical condition^4^. On paper, this approach promises reduced wait times, improved patient turnover, and increased patient satisfaction^5^. However, potential trade-offs must be recognized: are lower-acuity patients being prioritized at the expense of those with more urgent needs?

A key tool used in ED triage is the Emergency Severity Index (ESI), a widely accepted system that stratifies patients based on their acuity to determine the order in which they should be seen. The ESI categorizes patients into five levels, ranging from 1 (most acute) to 5 (least acute)^6^. Patients with ESI levels 1 and 2 require immediate evaluation and intervention classified as ESI 3 are considered urgent and can safely wait a short time for care. Patients with ESI levels 4 and 5, deemed non-urgent, can wait longer without significant risk^7^. Although the ESI system is designed to reflect both the severity of a patient’s condition and the anticipated use of ED resources, the actual assignment of an ESI level often relies on the triage staff’s clinical judgment, experience, and standardized criteria, such as vital signs^6^.

At first glance, the split-flow model is a win-win solution for patients and hospitals. Lower-acuity cases get quick treatment, freeing up resources for more severe cases^8^. Yet, our investigations of the Long Island Jewish Medical Center (LIJMC) split-flow model uncovered that up to 40% of lower-acuity patients were seen by providers before those with higher-acuity. This means that patients classified as more-acute based on the ESI sometimes wait longer for care than those with less-urgent conditions. Even within the higher-acuity group (ESI 1-3), the study found that less urgent cases (ESI 3) are sometimes seen before more critical ones (ESI 2)^2^. This inversion of care prioritization raises significant ethical and clinical concerns, particularly in emergency medicine, where every second counts for patients with serious, time-sensitive conditions^9^.

This study aimed to investigate whether prioritizing less-acute patients over more-acute ones influences outcomes such as patient disposition, including admission into the hospital vs. discharge from ED and the rates of 30- and 90-day return visits and 30-day admissions. We hypothesized that more-acute patients seen after a near-simultaneously-arriving less-acute patients are may have higher rates of return visits and increased hospital readmissions compared to more-acute patients seen before near-simultaneously-arriving less-acute patients on account of delays in care.

## MATERIALS AND METHODS

The data used in this analysis were obtained from our previous study^2^. This retrospective observational study assessed the impact of the split-flow model in the Emergency Department (ED) of Long Island Jewish Medical Center (LIJMC) on the order in which more-acute patients are seen compared with near-simultaneously-arriving less-acute patients, with acuity determined by use of the ESI at triage.

The Long Island Jewish Medical Center (LIJMC) is a 583-bed tertiary-care academic hospital serving a racially and socioeconomically diverse population. The adult ED sees approximately 100,000 patients per year and has an internal area designated for psychiatric patients.

After either walking in or arriving by ambulance, patients are triaged by nurses to either a 20-bed Fast Track area (for ESI levels 3, 4, and 5 patients), an acute care area (to which all ESI levels can be sent), or the psychiatry area in the ED. As part of the triage process, a brief chief complaint and vital signs are obtained, as well as blood sugar measurement and ECG evaluations, as indicated. The ED does not utilize a “provider-up-front” model^10^ whereby a provider assists with the triage process or rapidly screens a patient and orders studies or medications.

All ED treatment areas are staffed by patient care assistants (similar to medical assistants), ED Technicians (repurposed paramedics), nurses, and providers (Attending and resident physicians, physician assistants, and (rarely) nurse practitioner mid-level providers). Throughout the ED, laboratory or radiology studies are ordered only by providers after they have seen a patient and not by staff other than providers (eg, by a nurse operating under standardized procedure).

A convenience sample of 126 patient pairs aged ≥18 years was taken between April 24 and December 13, 2023. Patients arriving during any shift when a physician author (MR and DS) was present were included. Patient data was captured by research assistants during a wide variety of possible sampling times, given the variability in ED shiftwork (any time of day, evening, or night; weekdays and weeknights). Any pair of patients whose triage times were within 10 minutes of each other were selected and included as long as the provider sign-up time for each patient was after triage, so the ESI level was available for the provider to see, as that might affect how the provider prioritized which patient to see first. Patients in a pairing could have been triaged either in the same area (eg, both patients to Fast Track) or in different areas (eg, one patient to acute care, one patient to Fast Track). Patients were excluded if they were either directly sent to the psychiatric area of the ED after triage or if any of the authors were their Attending physicians. That is, patients were enrolled in the study during times when a physician author was on shift, but no study patients were attended to by either physician author.

The collected variables included the ESI level, the ED location to which the patient was triaged (either Fast Track vs. acute care area) and timestamps for triage and first-provider sign-up. Additionally, data were gathered including return visits at 30 and 90 days and patient disposition during the 30-day revisit. Patients admitted for observation or transfer were classified as admitted.

The patient pairings were divided into those in which the lower-acuity patient was seen first and those in which the higher-acuity patient was seen first. The study assessed the percentage of more-acute patients returning for follow-up visits within 30 and 90 days of their initial visit, as well as their readmission into the hospital and discharge from the ED rates within 30 days, comparing more-acute patients who were seen before vs. after the near-simultaneously-arriving less-acute patients.

A Students T-test was used to assess for statistically-significant differences in 30- and 90-day revisits and 30-day admissions. Statistical significance was established a priori at p <0.05.

This study received an exemption from the Northwell Health Institutional Review Board (IRB #23-0169).

## RESULTS

The study includes 126 patient pairs (252 patients in total). There were no statistically-significant differences in dispositional outcomes for more-acute patients based on the order in which they were seen. (Table 1).

**Table 1:** Disposition outcomes among more-vs. vs. less-acute patients being seen first by provider.

| Outcomes | % patients at index visit (N) | % admitted at index visit (N) | % Return within 30-Day visit (N) | % of patients who returned and admitted within 30 days (N) | % return within 90 days (N) |
| --- | --- | --- | --- | --- | --- |
| More-acute patients seen first | 60 (76) | 47 (36) | 10 (8) | 63 (5) | 20 (15) |
| Less-acute patients seen first | 40 (50) | 50 (25) | 22 (11) | 45 (5) | 26 (13) |
| P-value | 0.0285 | 0.2450 | 0.5029 | 0.8718 | 0.7109 |

## DISCUSSION

Our previous study on the split-flow model, comparing Fast-Track to main ED (acute care area), revealed that, approximately 40% of the time, less-acute patients were seen prior to near-simultaneously-arriving more-acute patients^2^. This goes against the core ethical principle of emergency medicine, which states that those with the greatest clinical need should be evaluated and treated first, raising potential ethical concerns ^11-13^. Such delays for more-acute patients could lead to adverse outcomes, including greater rates of hospital readmissions and 30- and 90-day ED return visits for more-acute patients.

Although our study found no significant differences in clinical outcomes based on the order in which patients were seen, the data shows important trends. Notably, when more-acute patients were seen first, 10% returned to the ED within 30 days, with 63% requiring hospital admission, and 20% of indexed patients returned within 90 days. However, when less-acute patients were seen first, the 30-day return rate for more-acute patients increased to 22%, with 45% needing hospital admission, and the 90-day return rate rose to 26%. This suggests that delays in treating more-acute patients might result in worse outcomes, possibly because of delayed care during their first ED visit or developed complications that required them to return.

Research has shown that the timeliness of care significantly impacts patient outcomes in emergency settings. For instance, timely administration of treatments such as analgesia and antibiotics is crucial for improving patient recovery and minimizing complications. Delayed analgesia is associated with worse pain management outcomes, while timely antibiotic administration for suspected sepsis can be life-saving ^14^. Additional adverse outcomes of long wait times for treatment include increased mortality, hospital-acquired infections, and higher readmission rates^15-18^. Instituting a patient flow policy and procedure that routinely (40% of the time) allows less-acute patients to be evaluated and treated prior to more-acute patients may unintentionally increase the risk of harm to more-acute patients, whose care is delayed as less-acute patients are seen first.

The ethical implications of these findings warrant careful consideration. While the split-flow model has demonstrated numerous benefits - such as decreased length of stay (LOS), improved patient flow (and, hence, increased revenue), and higher patient satisfaction - it may conflict with the principle of beneficence, which is the obligation to “do no harm”^19^. This ethical principle demands that healthcare providers prioritize the well-being of patients and ensure that care decisions do not unintentionally result in harm, particularly for those with higher-acuity.

The principle of beneficence requires continual reassessment of practices that, while operationally effective, may not fully benefit patients, particularly those with the greatest need. This principle requires us to continually reassess practices we assume benefit patients, acknowledging the possibility that we may be mistaken in our assumptions^19-20^. The observations that delays in care (eg, antibiotics in sepsis) are associated with worse clinical outcomes should prompt ED administrators to rethink their approach to balancing the needs of more-acute arrivals with those of less-acute patients, who still deserve to be seen in a timely manner.

While it is generally the goal of an ED to prioritize more-acute patients over less-acute ones, it is not always practical to ensure that every high-acuity patient is seen before lower-acuity patients. This challenge arises because, as fewer sick patients wait to be seen, additional higher-acuity patients may arrive, continuously pushing the lower-acuity patients further down the queue. ED administrators might consider several strategies to balance the need to see both higher and lower-acuity patients promptly: 1) Institutions could consider involving providers directly in the triage process (“provider-up-front”)^10^ to minimize the time between a patient’s arrival and their first contact with a provider, thus streamlining the process and expediting care for all acuity levels, 2) Instead of separating high- and low-acuity patients into different treatment areas, EDs could integrate patients of varying acuity into the same areas. This would allow providers to manage a mix of both patient types, helping to balance the urgency of care across the board. 3) EDs might establish a reasonable waiting time for low-acuity patients (e.g., two hours) before being seen. If this time limit is exceeded, a provider would be notified to address the patient unless they are actively attending to a more critical patient. 4) Under the Emergency Medical Treatment & Labor Act (EMTALA), EDs are required to conduct medical screening examinations (MSEs) to determine a patient’s level of severity. In practice, these exams often evolve into full assessments, even for minor complaints. However, in some states, including New York, providers, under law can, expedite the discharge of low-acuity patients to home or arrange rapid follow-up care at lower-acuity facilities, such as urgent care centers, potentially owned by the same organization as owns the ED^2^.

Incorporating these strategies into future efforts may help address the conflict between seeing lower-acuity patients quickly while ensuring higher-acuity patients are seen as soon as possible. This approach may reduce overall patient wait times and improve the efficiency of ED operations.

## Limitations

This study has several limitations. First, it was a single-center study; results may differ at other institutions, because LIJ ED’s triage patterns and patient flow may not reflect those in other EDs. Second, we used the timestamp of provider sign-up to indicate when a provider first saw the patient. However, sometimes a provider might sign up for a patient shortly after they are triaged to that provider’s area, without seeing the patient until the patient arrives in the provider’s area. In such circumstances, signing up for the patient indicates not the “time seen,” but, rather, the time the provider took responsibility for the patient. Third, as this was an exploratory study, sample sizes across comparison groups were small and, consequently, uneven, with some groups having especially-low numbers. This uneven distribution might have reduced our ability to detect true differences between groups.Finally, the relatively-small sample size (126 pairs) might have limited the ability to to detect true outcome differences to a degree of a-priori-stated statistical significance (p <0.05); had there been a larger sample size, the trend toward worse outcomes may have achieved statistical significance. We intend to conduct future, larger studies.

## CONCLUSION

While encouraging a triage system in which less-acute patients can frequently (nearly 40% in our study) be seen before more-acute patients presents advantages in terms of ED efficiency, this study suggests it may inadvertently lead to poorer outcomes for more-acute patients, particularly concerning revisit and admission rates. Healthcare institutions must carefully consider these findings when designing and implementing process innovations, ensuring that operational goals do not supersede the ethical imperative to prioritize the sickest patients.

## Data Availability

All data produced in the present study are available upon reasonable request to the authors.

## Acknowledgements

None.

